# Radiomic Analysis of Breast Thermal Images Using Thermalytix: A Multi-case, Multit-reader Study Against Manual Interpretation

**DOI:** 10.1101/2023.01.31.23285320

**Authors:** HV Ramprakash, Sudhakar Sampangi, Shylaja Swami, Siva Teja Kakileti, Geetha Manjunath, Sathiakar Collison

## Abstract

**Objective:** To evaluate the performance of Thermalytix, an artificial intelligence-enhanced breast thermal imaging analysis software, against unaided manual interpretation of thermographic images.

**Methods:** In this multi-reader study, thermal imaging data of 258 symptomatic participants from a previous clinical trial were used. These images were independently manually interpreted by 3 senior trained breast radiologists. The same images were independently evaluated by Thermalytix, which uses sophisticated machine learning analysis of thermal/ vascular radiomic parameters to generate a risk score predictive of cancer . The results of manual interpretation and Thermalytix were compared with reference standard based on standard of care (combination of mammography, ultrasound and histopathology), to determine sensitivity, specificity, positive predictive value (PPV), and negative predictive value (NPV) and area under receiver operating characteristic curves (AUROC).

**Results:** Thermalytix obtained showed a sensitivity and specificity of 95.2% (90% confidence interval (CI), 90.0- 100.0) and 66.7% (CI 60.1-73.3); the NPV and PPV were 97.7% (CI 95.2%-100.3%) and 58.3% (CI 48.5%-68.2%). The (sensitivity, specificity, NPV, PPV) obtained by Reader 1, Reader 2 and Reader 3 were (60.3%, 81.5%, 51.4%, 86.4%), (74.6%,50.8%, 86.1%, 32.9%) and (71.4%, 63.8%, 87.2%, 38.5%), respectively. The AUROC of Thermalytix was 0.85, 13.7% greater than manual interpretation.

**Conclusion:** Thermalytix demonstrated good clinical performance with 25% higher accuracy than manual interpretation of thermal images. Thermalytix may alleviate the known subjectivity in thennography thereby improving its performance.

## Introduction

Infrared thermal imaging has been used in medicine since the early 1960s [1]. However, due to technological limitations such as low image resolution and poor temperature sensitivity, early imaging systems delivered sub-optimal performance [2]. In contrast, modern high-resolution thermal cameras represent a significant leap forward. These systems use micro-electromechanical technologies, large 2D focal plane array detectors, vanadium oxide microbolometers, and on-chip image processing to enhance thermal sensitivity and spatial resolution, all without requiring active cooling [3]. These cameras are compact, lightweight, and affordable due to advances in silicon wafer fabrication [4]. These substantial improvements, along with a growing recognition of the benefits of non-invasive, non-ionizing imaging techniques, have led to a renewed interest in medical thermography across several clinical applications [2].

One such application is for the detection of breast cancer. The rationale for using thermal imaging in breast cancer detection is grounded in the physiological changes that accompany malignancy. Cancerous tissues exhibit high metabolic rates and increased angiogenesis-both hallmarks of cancer [5] and are characterised by increased localized blood flow, vasodilation, increased vessel tortuosity, and the formation of new blood vessels. The resultant increased regional heat production is transferred to the skin surface and can be captured using high-resolution thermal imaging systems [6].

Although the physiological basis for thermal imaging in breast cancer is sound, early clinical applications produced highly variable results. One limitation was the poor thermal sensitivity of earlier cameras. However, an equally important shortcoming was the reliance on visual interpretation of thermal images. While the sensor in thermal cameras do measure actual temperature values, by default, these are typically converted into colorized pseudo-images using palettes such as rainbow, iron, or grayscale [7]. This conversion depends on the use of a predefined temperature range, introducing variability in the displayed image. Consequently, the same thermal data could appear markedly different depending on the selected palette and scale. Moreover, human visual perception is inherently limited for the task of discriminating between subtle color gradients, making it difficult for even expert readers to detect subtle thermal anomalies [7]. This subjectivity, compounded by the lack of standardised protocol for image capture and the fact that some benign conditions also exhibit increased metabolic heat, often led to false positives or missed diagnoses [8].

Manual interpretation of thermal images, therefore, required experienced readers but remained inherently qualitative and unstructured. All these issues, along with inconsistent imaging protocols and inadequate device sensitivity, were key factors that led to the unsatisfactory diagnostic accuracy reported in earlier large-scale evaluations such as the Breast Cancer Detection Demonstration Project (BCDDP) [9]. Despite these challenges, the U.S. Food and Drug Administration acknowledged the potential value of thermography and approved it in 1982 as an adjunctive imaging tool for breast cancer detection. This recognition was based on its value as a functional imaging modality that could complement anatomical imaging techniques.

However, even with the superior sensitivity and resolution of current thermal cameras-often detecting temperature changes as small as 0.01 to 0.05 °C and spatial features under 2 mm-it remains difficult for the human eye to interpret nuanced thermal variations. The cognitive load involved in deciphering complex color maps, combined with the absence of a structured scoring system, continues to limit the diagnostic utility of manual interpretation of breast thermal images.

In the last decade, advances in radiomics and machine learning have introduced a paradigm shift in image interpretation, which have been applied towards improving the analysis of breast thermograms. Rather than relying on subjective human observation, these approaches extract quantifiable features-such as pixel-level temperature patterns and vascular distributions-allowing for standardized, objective interpretation [10]. These artificial intelligence (AI)-based algorithms may substantially reduce inter-reader variability and improve diagnostic accuracy [11,12].

One such AI-based system is Thermalytix, which uses radiomics-based machine learning algorithms to automatically generate a score predictive of the presence of malignancy. The platform analyzes medically interpretable thermal and vascular features that reflect underlying metabolic and vascular dynamics [13]. This is conceptually similar to the application of radiomics in mammography, where machine learning is used to extract, characterize, and classify image features to support diagnosis [14].

In this study, we compare the performance of Thermalytix, an automated breast thermography interpretation platform, against manual interpretation by expert thermologists. We use data from a previously published prospective multicenter clinical trial and evaluate whether AI-based analysis can offer more objective and reliable diagnostic performance than manual interpretation of breast thermal images.

## METHODOLOGY

### Study Population

We retrospectively analyzed data from a previously published, Institutional Review Board-approved, prospective multicenter clinical trial (CTRI/2017/l 0/010115) [15], conducted between September 2017 and July 2018 at two clinical sites in Bangalore, India: Narayana Hrudayalaya (NH) and HealthCare Global (HCG). In the original observational study, all participants underwent breast thermal imaging with analysis using the Thermalytix software. Each participant also underwent conventional imaging involving mammography and/or breast ultrasound as clinically indicated, with any suspicious findings subsequently confirmed through histopathology.

Thermal imaging was performed prior to other imaging modalities to avoid potential artifacts from breast compression (mammography) or contact gel application (ultrasound). Ethics committee approval was obtained at both sites for data collection and analysis in the prospective setting. All participants provided written informed consent at the time of enrollment. The retrospective analysis presented here was conducted as a secondary analysis using the latest version of the Thermalytix AI algorithm, after appropriate data anonymization. All procedures were carried out in compliance with institutional guidelines and applicable regulations.

Of the 326 women originally enrolled in the study, 68 were excluded due to incomplete data, as detailed in our prior publication [15]. The remaining 258 women, all of whom had a confirmed diagnosis based on standard imaging and/or histopathology, were included in the present analysis. Of these, 204 participants (79.1%) were recruited from NH, and 54 (20.9%) from HCG.

### Study Design

This was a multi-reader diagnostic performance study. Thermal images obtained from both participating sites were uploaded to the Thermalytix software platform, which automatically analyzed the images using proprietary machine learning algorithms to generate an objective malignancy risk score. This constituted the Thermalytix test output group.

For manual interpretation, the same thermal images-without any AI-derived outputs-were independently reviewed by three experienced readers. One was a senior breast radiologist and certified manual thermologist with over 15 years of experience in breast thermography. The other two were senior breast radiologists who had completed a standardized training program in thermographic interpretation prior to participation in this study. These three constituted the manual thermography reader group.

All manual readers were blinded to the final clinical diagnoses and to the Thermalytix output scores to ensure an unbiased assessment (Figure 1).

**Figure 1.**
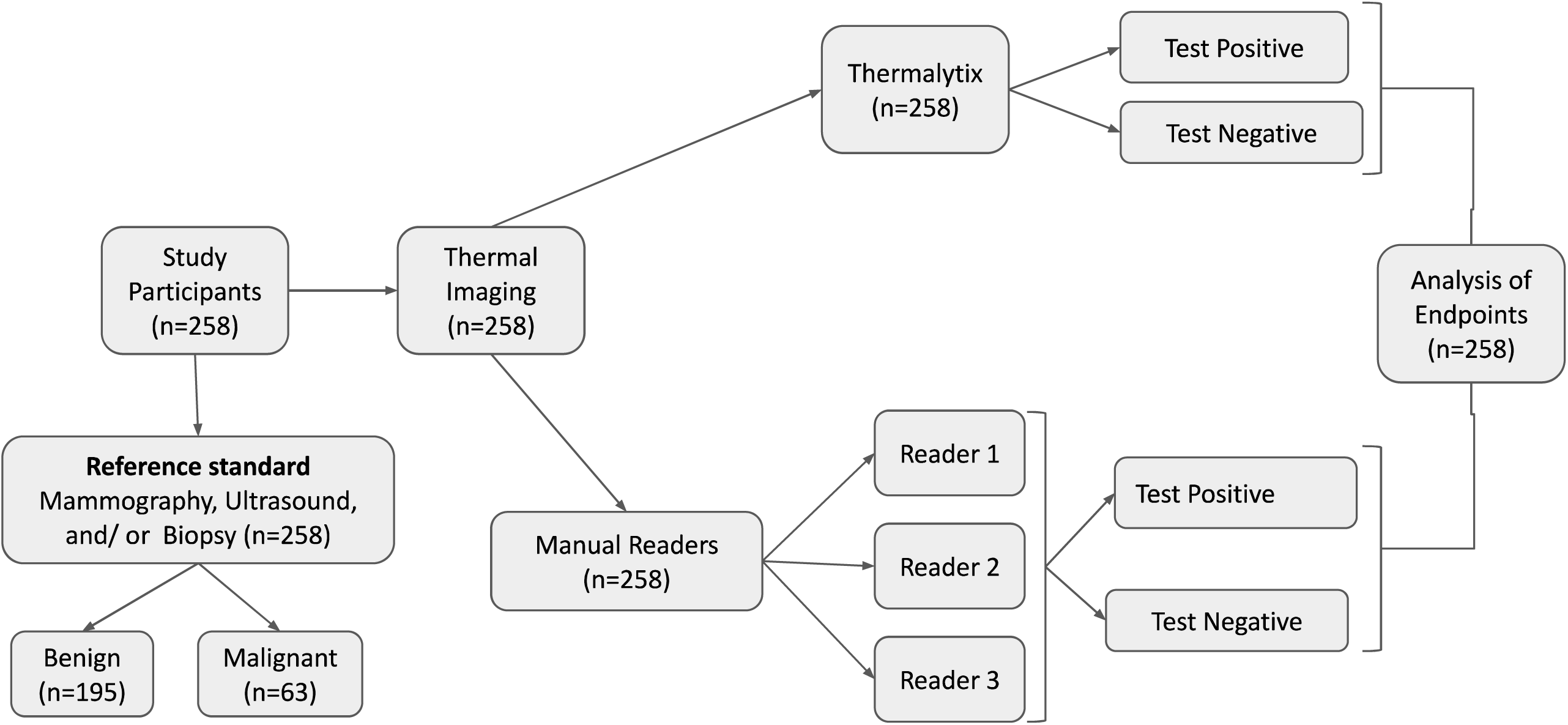
Diagram of the flow of the study protocol.

### Thermal Imaging Protocol

Thermal imaging was performed under controlled conditions. An air cooling system was used. Participants were asked to disrobe from the waist upwards and were allowed to equilibrate with the ambient temperature for 5 to 10 minutes prior to imaging. Thermal imaging was performed with the participant sitting on a rotating stool with the thermal camera about one metre away. At the time of imaging, the participants raised their arms above their heads, and five thermal images were captured at the following positions: frontal (0°), left oblique (45°), left lateral (90°), right oblique (-45°), and right lateral (-90°). This protocol ensured comprehensive thermal coverage of the breast surface (16).

### Technical specifications of the thermal cameras

Thermal imaging was performed using two digital infrared cameras from FLIR Systems (Wilsonville, OR, USA): the FLIR T650SC and the FLIR A3 l 5. The FLIR T650SC provides a spatial resolution of 640 x 480 pixels (307,200 pixels per scan) with a thermal sensitivity of< 0.02 °C, while the FLIR A315 offers a resolution of 320 x 240 pixels (76,800 pixels per scan) and a thermal sensitivity of< 0.05 °C..

### Manual Thermographic Assessment

For each participant, the five thermal images captured during imaging were provided to the manual readers, who independently assessed the presence or absence of findings suspicious for malignancy. The readers conducted a visual evaluation of the images without any computational assistance, using the Thermobiological Grading System as outlined by the American Academy of Thermology (AAT) guidelines [16]. This system incorporates a combination of qualitative and quantitative thermographic indicators that are suggestive of suspicious lesions.

Each participant’s thermal images were graded on a scale from O to 5, where higher grades indicate increasing suspicion of malignancy. In accordance with AAT recommendations, grades 3, 4, and 5 were considered test-positive for potential malignancy under manual interpretation.

### Computer-aided interpretation with Thermalytix

Thermalytix is an artificial intelligence-based, computer-aided diagnostic (CAD) software designed to automatically interpret breast thermal images. It utilizes a suite of machine learning algorithms to extract radiomic features from thermograms and generate a malignancy risk score. The output includes both a quantitative likelihood of malignancy and annotated visualizations highlighting suspicious thermal patterns (Figure 2).

**Figure 2:**
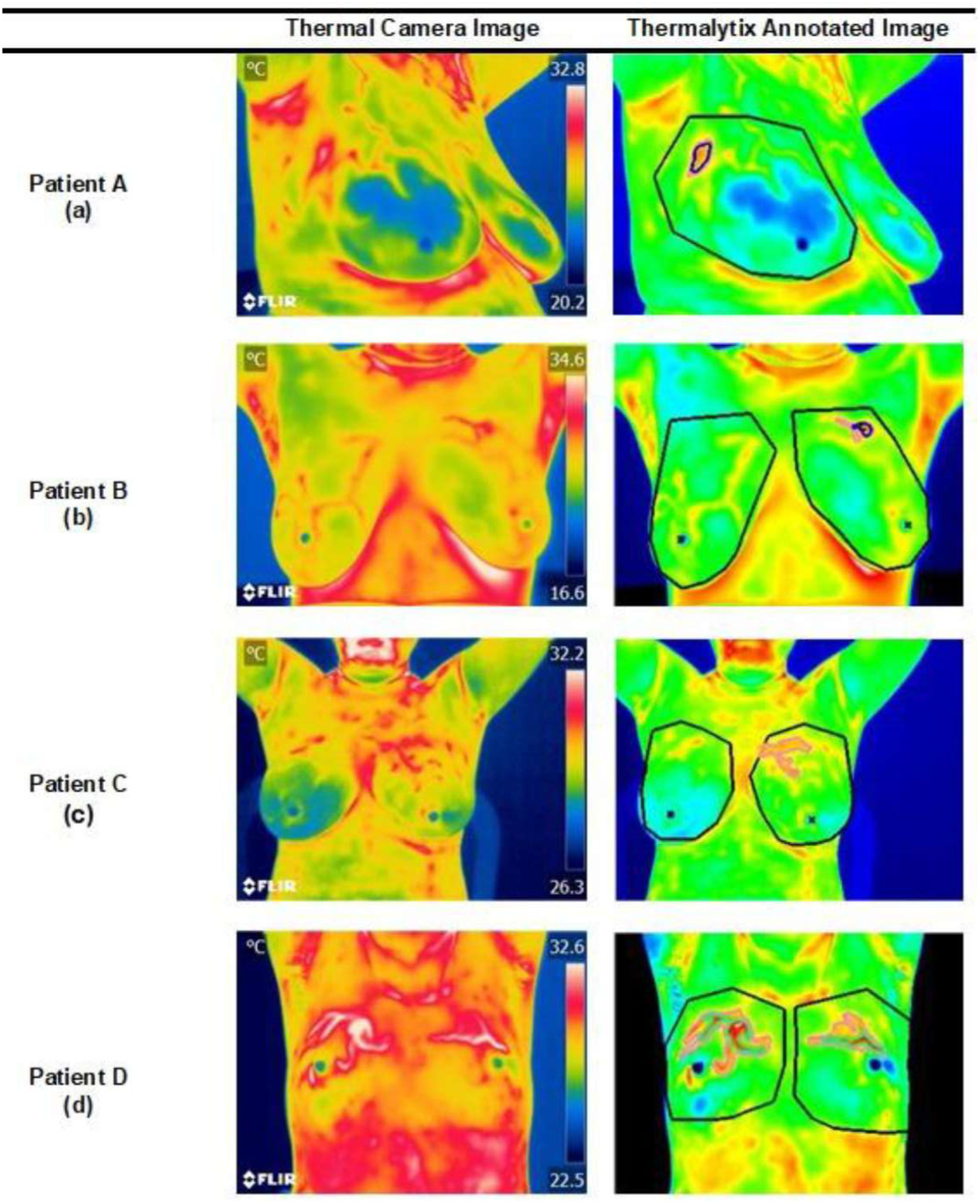
Case Studies showing raw thermal images alongside Thermalytix annotated images- (a) True positive for Thermalytix and False negative for all 3 manual readers, (b) True positive for Thermalytix and False negative for all 3 readers, (c) True negative for Thermalytix but False positive for 2 readers, (d) False negative ofThermalytix and True positive for 2 readers.

In the first step, automated image quality assessment is performed using pre-trained AI models, which evaluate parameters such as focus, view angle, and cooling adequacy to ensure diagnostic image integrity [Figure 3]. Following this, the images undergo preprocessing, wherein contrast enhancement is applied by auto-selecting minimum and maximum temperature thresholds. This step ensures consistent visualization of regions with elevated thermal activity across different patient images.

**Figure 3:**
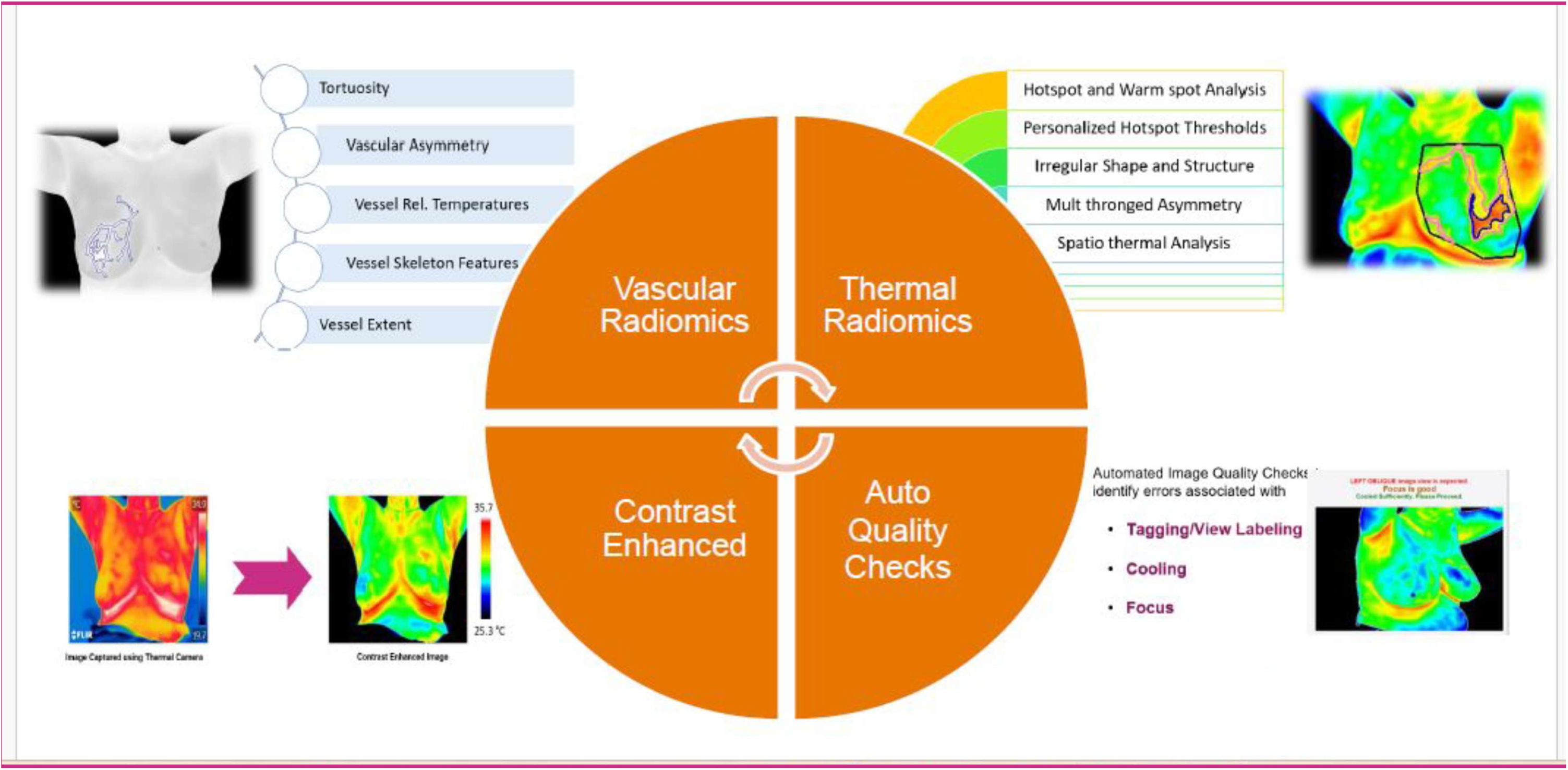
Schematic representation of the different stages where Thermalytix uses artificial intelligence and machine learning based computational algorithms towards its analysis. Please read the text for further details.

Subsequently, a dedicated AI model segments the breast regions from the thermal images. From these segmented regions, the system extracts three distinct sets of radiomic features: hotspot radiomics, vascular radiomics, and areolar radiomics, each capturing different functional aspects of thermal and vascular physiology [13,17].

- Hotspot radiomics characterize regions of increased thermal activity. Features such as shape, size, symmetry, temperature intensity, and spatial distribution are extracted from identified hot and warm spots [19].
- Vascular radiomics focus on detecting vascular structures and their symmetry. From these, 17 radiomic features are derived, including vessel count, branching pattern, mean calibre, symmetry index, and relative temperature metrics [18].
- Areolar radiomics assess thermal activity specifically within the areolar region, capturing asymmetries and local temperature deviations.

Each of the three radiomic categories is processed by a dedicated, pretrained machine learning classifier, which outputs independent scores: hotspot score, vascular score, and areolar score. These scores are subsequently integrated, along with relevant clinical parameters, into an ensemble classifier to compute the final Thermalytix B-score. A B-score greater than 3 is considered test-positive, flagging the case for further clinical evaluation. The system is intended to assist clinicians by objectively identifying high-risk thennographic patterns that warrant additional diagnostic investigations.

For more details of the processes involved in the artificial intelligence based analysis, the readers are directed to the methods section of the recent publication of Adapa et al [19].

### Statistical analysis

All statistical analyses were performed using IBM SPSS Statistics, Version 26.0 (IBM Corp., Armonk, NY, USA). Diagnostic performance metrics including sensitivity, specificity, positive predictive value (PPV), and negative predictive value (NPV) were calculated for both the Thermalytix system and manual thermographic interpretation, using the final clinical diagnosis as the reference standard. Receiver operating characteristic (ROC) curves were constructed for both methods to assess performance across various thresholds. The area under the ROC curve (AUROC) was computed to quantify overall diagnostic accuracy. The AUROC is mathematically equivalent to the Wilcoxon rank-sum (Mann-Whitney U) test statistic and reflects the probability that a randomly selected positive case will have a higher test score than a negative case.

Inter-reader agreement among the three manual interpreters was assessed using the kappa (K) statistic. Both Cohen’s kappa (for pairwise agreement) and Pleiss’ multirater kappa (for overall agreement across three readers) were computed to evaluate consistency in manual thermographic assessment.

## Results

Of the 258 women included in the study, 63 participants (24.4%) were classified as disease-positive, based on the reference standard. This included 33 malignancies from the Narayana Hrudayalaya (NH) site and 30 malignancies from the HealthCare Global (HCG) site. The remaining 195 women (75.6%) were considered disease-negative. Diagnostic performance of Thermalytix and manual thermography interpretation was evaluated against this ground truth.

The Thermalytix score yielded a sensitivity of 95.2% (90% CI: 90.0%-100.5%), specificity of 66.7% (90% CI: 60.0%-73.3%), positive predictive value (PPV) of 48.0% (90% CI: 39.2%-56.8%), and negative predictive value (NPV) of 97.7% (90% CI: 95.2%-100.3%).

For manual thermographic interpretation from three readers, the pooled diagnostic performance was: sensitivity of 68.8% (90% CI: 57.5%-80.1 %), specificity of 65.1% (90% CI: 58.7%-71.5%), PPV of 40.9% (90% CI: 31.6%-50.2%), and NPV of 86.6% (90% CI: 80.9%-92.2%). Detailed performance metrics for each reader are presented in Table 1.

Inter-reader agreement for manual interpretation was assessed using Cohen’s kappa statistic, which indicated only fair to moderate agreement among the three readers. These results are summarized in Table 2.

Receiver operating characteristic (ROC) analysis revealed an AUROC of 0.85 for the Thermalytix B-score. This was 12% higher than that of Reader 1 (AUROC = 0.73), 14% higher than Reader 2 (AUROC = 0.69), and 13% higher than Reader 3 (AUROC = 0.72) (Figure 4).

**Figure 4.**
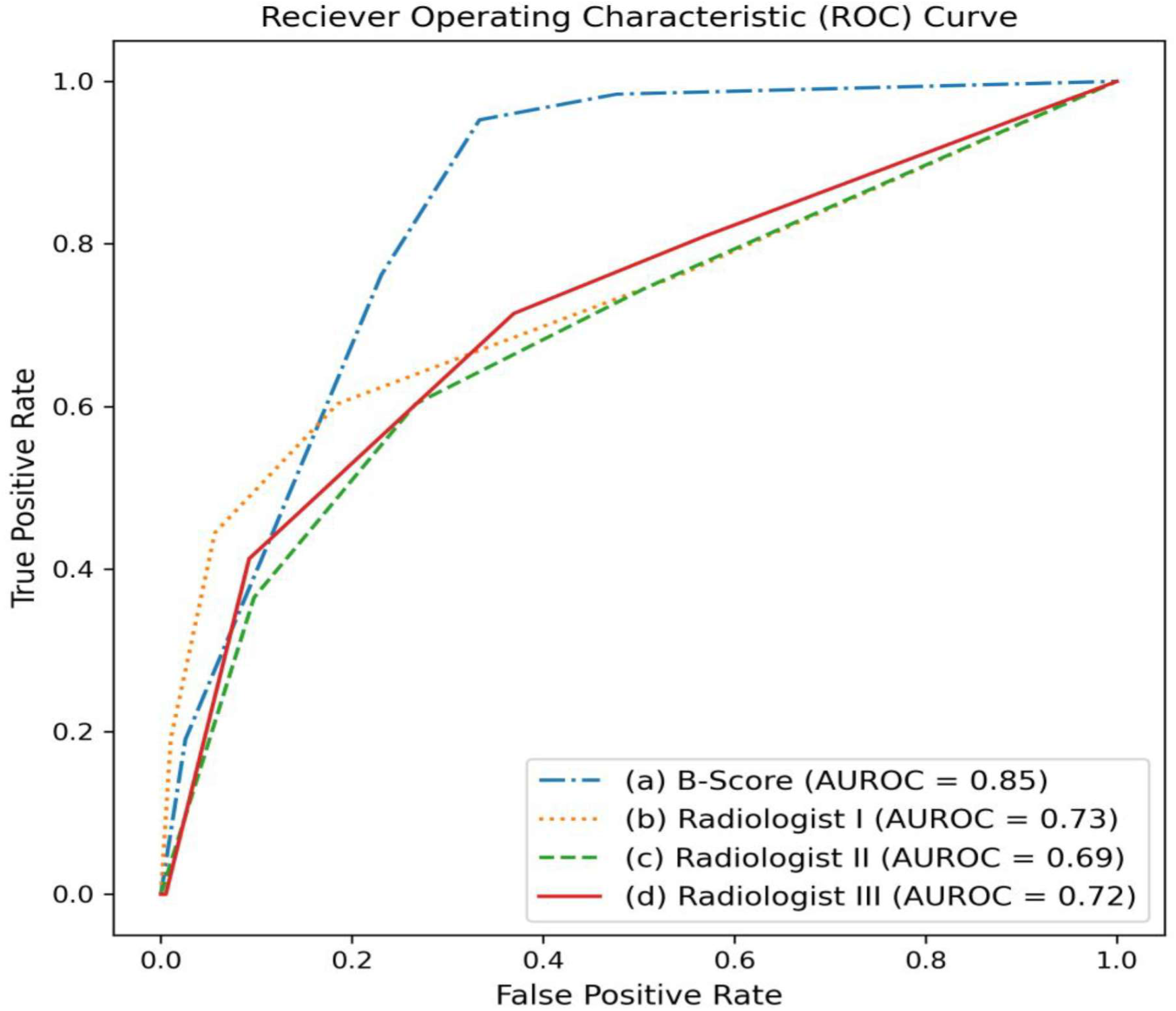
ROC curve demonstrating the separation in performance between Thermalytix and manual interpretation of thermography.

## Discussion

This multi-reader, multi-case study evaluated the performance of Thermalytix, an AI-based automated breast thermal image interpretation system, as compared with manual interpretation of the same images. In this study, Thermalytix output out-performed manual interpretation of breast thermography, with a 25% improvement in sensitivity and an AUROC that was 13.7% higher than the average AUROC obtained from manual readers.

Modern infrared thermal cameras offer remarkable improvements in both thermal sensitivity and spatial resolution, enabling pixel-level heat mapping with precise temperature measurement for each pixel. These advancements have markedly improved the quality and interpretability of breast thermal imaging, making it more clinically viable today than it was in previous decades. In our study manual interpretation still resulted in only moderate performance, with a mean sensitivity of 68.8%, specificity of 65.1%, PPV of 40.9%, and NPV of86.6%, underscoring the inherent limitations of human visual assessment, particularly in detecting subtle or asymmetric thermal patterns.

In the past, the Breast Cancer Detection Demonstration Project (BCDDP), a landmark trial in the 1970s, reported suboptimal performance of thermography in breast cancer detection [9], leading to its abandonment as a viable diagnostic tool. Key reasons included low thermal camera sensitivity, lack of standardized imaging protocols, and a reliance on subjective, unstructured manual interpretation. Interpretation at the time involved black-and-white imaging, vastly different from the high-resolution, color-mapped outputs available with today’s thermal systems [20,21]. Even then, practitioners acknowledged that thermal image quality was crucial and required skilled technicians, and interpretation was prone to considerable inter-reader variability.

Today, these limitations have been largely addressed. AI systems such as Thermalytix eliminate subjectivity by leveraging quantitative radiomic features and machine learning-based scoring algorithms [Figure 3]. Similar AI based techniques have proven useful in other domains of medical imaging, such as mammography, where it has successfully detected malignancies up to five years before conventional diagnosis [22]. Similarly as demonstrated here, for breast thermal imaging, AI can detect subtle variations that are imperceptible to the human eye and identify complex vascular or metabolic patterns that correlate with early malignancy [6,11,12,21].

In our study, Thermalytix’s AUROC of 0.85 was superior to all three manual readers. These findings are consistent with those from a larger prospective study involving 470 women, in which Thermalytix was shown to be non-inferior to mammography in sensitivity [23]. Bansal et al. [24] also reported sensitivity of95.24% and specificity of88.58% when evaluating Thermalytix.

Importantly, the analysis of Thermalytix uses features that are medically interpretable as the outputs are derived from radiomic analysis of hotspot and vascular thermal features. The system generates scores that can be understood even by non-specialist readers, making it highly adaptable in diverse clinical settings. Further, and our study showed low inter-reader agreement in manual interpretation, with Cohen’s and Fleiss’ kappa values reflecting only fair to moderate agreement (the commonly accepted threshold (K > 0.60) for diagnostic reliability [25]).

Thermalytix also incorporates automated quality control algorithms that validate image focus, patient positioning, and cooling compliance at the time of acquisition. This ensures that the thermal images meet diagnostic standards regardless of technician experience, thereby removing one of the longstanding barriers to the broader adoption of thermography [Figure 3]

Earlier efforts to integrate computer analysis with thermographic imaging used relatively simpler computer analytic tools.

In 1977, Negin et al. [26] developed a basic computer tool that showed a 4-7% improvement over manual classification. In 1999, Wiecek et al. [27] used MATLAB-based thermal signature calculations and wavelet-transformed parameters to detect biopsy-confirmed lesions. The Sentinel BreastScan, a U.S. FDA-approved device, was studied by Wishart et al. [28], who analyzed results using manual, AI (neural network), and software interpretations, finding that the Al’s sensitivity (48%) was low but the computer software’s performance (70%) closely approached expert manual review (78%).

In 2008, Arora et al. [29] examined 92 women with prior suspicious mammograms and reported Sentinel BreastScan sensitivity of 97% and NPV of 82%. Nair et al. [30] evaluated 180 women using the NoTouch BreastScan and reported AI sensitivity of 88.24%, specificity of 70.52%, NPV of 87.01%, and PPV of 72.82%. In contrast, mammography achieved higher specificity and NPV. Umadevi et al. [31] developed the ITBIC system that captured three thermal images and created simplified thermograms, achieving sensitivity of 66.7%, specificity of 97.7%, PPV of 80%, and NPV of 95.6%. Similarly, Acharya et al. [32] used a support vector machine on 50 thermal images, reporting sensitivity of 85.71% and specificity of 90.48%.

While this study demonstrates promising results, it has certain limitations. The analysis was retrospective, and the cohort was drawn from two clinical sites in southern India, which may limit generalizability. Additionally, the study lacked longitudinal follow-up to assess interval cancers or long-term outcomes. Future work should focus on prospective validation across diverse ethinic populations

## Conclusion

Recent advancements in infrared thermal imaging technology, when combined with state-of-the-art machine learning platforms such as Thermalytix, offer significantly improved performance over manual thermographic interpretation for breast cancer detection. By leveraging novel radiomic features, Thermalytix enables automated, interpretable, and quantitative analysis of breast thermal images, reducing subjectivity and inter-reader variability.

## Author contributions

HVR, SK and GM and H.K. conceived the study and methodology; HVR, SS (author 2), SS (author 3) and SK supervised the conduct of the experiment; SC was only involved in the preparation of the manuscript. All the authors equally contributed to review, and editing of the manuscript and in the verification of the underlying data reported in the manuscript. All authors have read and approved the manuscript.

## Data availability

Statistical and demographic data that underlie the results reported in this article after de-identification will be shared upon request directly to the corresponding author. The data will be available beginning 3 months and ending 2 years following article publication. Researchers who provide a methodologically sound proposal should approach the corresponding author; to gain access, data requestors will need to sign a data access agreement. Ethical and legal implications of data sharing will be considered and the decision to share data will be based on the outcome of this review.

## Code availability

The underlying code for this study and training/validation datasets is not publicly available for proprietary reasons.

