## Supplementary material for "Radiomic Analysis of Breast Thermal Images Using Thermalytix: A Multi-case, Multit-reader Study Against Manual Interpretation": Tables: JAMA tables.pdf

**Table 1.** Sensitivity, Specificity, Positive Predictive Value and Negative Predictive Value of Thermalytix and Manual Thermography.

| | **Sensitivity** | **Specificity** | **PPV** | **NPV** |
|---|---|---|---|---|
| B-Score with automated Thermalytix | 95.2% (90.0%-100%) | 66.7% (60.0%-73.2%) | 48.0% (39.2%-56.8%) | 97.7% (95.2%-100%) |
| **Manual Thermography** | | | | |
| Reader 1 | 60.3% (48.2%-72.3%) | 81.5% (76.0%-87.0%) | 51.4% (41.8%-62.7%) | 86.4% (82.3%-91.3%) |
| Reader 2 | 74.6% (63.9%-85.4%) | 50.8% (43.8%-58.7%) | 32.9% (25.2%-41.0%) | 86.1% (79.8%-92.4%) |
| Reader 3 | 71.4% (60.3%-83.4%) | 63.1% (56.3%-70.8%) | 38.5% (29.6%-47.3%) | 87.2% (81.7%-92.7%) |

The 90% confidence interval is provided in brackets

**Table 2:** Kappa scores

| Cohen Kappa | | |
|---|---|---|
| **Readers selected** | **Raw Scores** | **Binary outcome [positive/ negative]** |
| Reader 1 vs Reader 2 | 0.234 | 0.384 |
| Reader 1 vs Reader 3 | 0.247 | 0.395 |
| Reader 2 vs Reader 3 | 0.303 | 0.594 |
| **Fleiss Kappa** | | |
| For all 3 readers | 0.248 | 0.443 |

Kappa is interpreted as follows: 0- no agreement; 0.01–0.20 slight agreement; 0.21–0.40 fair agreement; 0.41– 0.60 moderate agreement; 0.61–0.80 substantial agreement; 0.81–1.00 almost perfect agreement.
